# Respiratory Virus Circulation during the First Year of the COVID-19 Pandemic in the Household Influenza Vaccine Evaluation (HIVE) Cohort

**DOI:** 10.1101/2022.12.08.22283268

**Authors:** Sydney R. Fine, Latifa A. Bazzi, Amy P. Callear, Joshua G. Petrie, Ryan E. Malosh, Joshua E. Tucker, Matthew Smith, Jessica Ibiebele, Adrian McDermott, Melissa A. Rolfes, Arnold S. Monto, Emily T. Martin

## Abstract

**Background:** The annual reappearance of respiratory viruses has been recognized for decades. The onset of the COVID-19 pandemic altered typical respiratory virus transmission patterns. COVID-19 mitigation measures taken during the pandemic were targeted at SARS-CoV-2 respiratory transmission and thus broadly impacted the burden of acute respiratory illnesses (ARIs), in general.

**Methods:** We used the longitudinal Household Influenza Vaccine Evaluation (HIVE) cohort of households in southeast Michigan to characterize mitigation strategy adherence, respiratory illness burden, and the circulation of 15 respiratory viruses during the COVID-19 pandemic determined by RT-PCR of respiratory specimens collected at illness onset. Study participants were surveyed twice during the study period (March 1, 2020, to June 30, 2021), and serologic specimens were collected for antibody measurement by electrochemiluminescence immunoassay. Incidence rates of ARI reports and virus detections were calculated and compared using incidence rate ratios for the study period and a pre-pandemic period of similar length.

**Results:** Overall, 437 participants reported a total of 772 ARIs and 329 specimens (42.6%) had respiratory viruses detected. Rhinoviruses were the most frequently detected organism, but seasonal coronaviruses—excluding SARS-CoV-2—were also common. Illness reports and percent positivity were lowest from May to August 2020, when mitigation measures were most stringent. Study participants were more adherent to mitigation measures in the first survey compared with the second survey. Supplemental serology surveillance identified 5.3% seropositivity for SARS-CoV-2 in summer 2020; 3.0% between fall 2020 and winter 2021; and 11.3% in spring 2021. Compared to a pre-pandemic period of similar length, the incidence rate of total reported ARIs for the study period was 50% lower (95% CI: 0.5, 0.6; p<0.001) than the incidence rate from March 1, 2016, to June 30, 2017.

**Conclusions:** The burden of ARI in the HIVE cohort during the COVID-19 pandemic fluctuated, with declines occurring concurrently with the widespread use of public health measures. It is notable, however, that rhinovirus and seasonal coronaviruses continued to circulate even as influenza and SARS-CoV-2 circulation was low.

## BACKGROUND

The annual reappearance of influenza has been recognized for decades through clinical identification and systematic surveillance of influenza-like illnesses.^1,2^ The addition of molecular-based testing to clinical and public health surveillance systems has established patterns of seasonality for non-influenza viruses, as well. While specific timing and intensity can vary, broader patterns for specific viruses are remarkably predictable from year to year, with respiratory syncytial virus, influenza, coronaviruses, and human metapneumovirus increasing in the winter and parainfluenza increasing in the fall and/or spring.^3^ Rhinoviruses are detected year-round, but increases in spring and fall are typical.^4,5^ Many hypotheses exist to explain virus-specific patterns of surge at different times of the year, including virologic, environmental, and social factors involved in transmission, and some of these potential explanations have been examined experimentally.^6–8^ Before the COVID-19 pandemic, few variations from seasonal patterns were observed, with the important exception of the occurrence of out-of-season influenza transmission during the 2009 influenza pandemic.^9^

The start of the COVID-19 pandemic in the United States in March 2020 came at the end of the tenth year of the HIVE (Household Influenza Vaccine Evaluation) study. During the previous ten years of HIVE surveillance, the occurrence of influenza and other respiratory viruses (including the seasonal coronaviruses) was followed in a household cohort. Year-round acute respiratory illness (ARI) surveillance began in 2015.^10–13^ The study conducted active surveillance for ARIs among cohort enrollees during the COVID-19 pandemic once new strategies to collect specimens were implemented according to SARS-CoV-2 transmission-control restrictions.^14^ Here, we report changes to the circulation of respiratory viruses throughout multiple early waves of the pandemic in a cohort of households reporting high levels of COVID-19 mitigation practices with repeated serosurveys for the acquisition of SARS-CoV-2 antibodies.

## METHODS

### Study Population

HIVE is a longitudinal cohort that has followed approximately 300-400 households per year in southeastern Michigan since 2011 using active surveillance for ARI. Methods for recruitment of participants and participation in the HIVE study have been previously described.^10^ Eligible households must have ≥3 persons living at the same address with at least one child aged <10 years at the time of initial enrollment. In mid-2020, eligibility was expanded to those with at least one child aged <21 years. Data included in this analysis represent the period from March 1, 2020, to June 30, 2021, to capture individuals enrolled during the COVID-19 pandemic. We retained households throughout the study period even if children aged out of the original eligibility criteria. Participants were removed from the study follow-up only if they chose to withdraw. HIVE is approved by the institutional review board at the University of Michigan Medical School; a reliance agreement was established with CDC to cover this work.

### ARI Surveillance and Laboratory Confirmation of Infection

Households received weekly e-mail reminders to report ARIs defined as the presence of ≥2 of the following symptoms: cough, fever/feverishness, nasal congestion, chills, headache, body aches, or sore throat for those ≥3 years old, or cough, fever/feverishness, nasal congestion/runny nose, trouble breathing, fussiness/irritability, decreased appetite, or fatigue for those <3 years old. For those who met the case definition, nasal swabs were obtained at symptom onset (<7 days post-onset) by parents or self-collection. Specimens were tested by multiplex reverse-transcription polymerase chain reaction RT-PCR (Fast Track Diagnostics, Respiratory Pathogens 33 kit [FTD-2P.3-64-RUO], Siemens Healthineers, Malvern, PA) for rhinovirus (RV), adenovirus (AdV), human coronaviruses (NL63, OC43, HKU1, and 229E), enterovirus (EV), human bocavirus (HBoV), human metapneumovirus (hMPV), parainfluenza virus (1, 2, 3, and 4) (HPIV), respiratory syncytial virus (RSV), and parechovirus (PeV). SARS-CoV-2 was detected by RT-PCR using CDC Research-Use Only protocols. Laboratory testing was performed at the University of Michigan School of Public Health.

### Serologic Testing

Enrollees participated in blood draws during three serosurveillance periods: summer 2020 (range: 7/17/2020-8/31/2020), fall-winter 2020 (range: 9/1/2020-2/3/2021), and spring 2021 (range: 3/9/2021-4/30/2021). Serum specimens were tested by electrochemiluminescence immunoassay (ECLIA) (Meso Scale Discovery, Rockville, Maryland) to measure antibody levels against SARS-CoV-2 spike protein, SARS-CoV-2 spike protein receptor-binding domain (RBD), and SARS-CoV-2 nucleocapsid (N) protein using methods previously described.^11^ A result of 23.5 IU/mL N protein, 14.1 IU/mL RBD, and 10.8 IU/mL spike protein was considered positive. For unvaccinated individuals, a positive result against any target was considered to indicate prior infection. Once an individual was vaccinated, only a positive result for the N protein target was considered indicative of prior infection.

### Mitigation Survey

Adults from participating households were surveyed from July-August 2020 and May-July 2021 on COVID-19 mitigation practices. The first survey included questions to assess changes in work and school attendance outside of the home, sources of trusted information, essential worker status, and common mitigation practices (e.g., masking, distancing, hand hygiene, etc.) (Supplemental Materials). The second survey included additional questions regarding COVID-19 vaccination (Supplemental Materials).

### Statistical Analysis

ARI reports were summarized for the study population by week and age group from March 1, 2020, through June 30, 2021. Incidence rates (IR) of ARI reports and virus detections were calculated per 100 person-years for the study period and for a pre-pandemic period of similar length (March 1, 2016, to June 30, 2017); these rates were compared using incidence rate ratios (IRR) with 95% confidence intervals (CI). The detection of each respiratory virus as a proportion of reported ARIs was determined overall and compared by age group (<5, 6-11, 12-17, 18-49, ≥50 years). For analytic purposes, *child* is defined as age <18 years. An epidemic curve for the study period was generated by plotting ARI reports by RT-PCR test result. Reported COVID-19 cases in Michigan were obtained from the MI Safe Start Map (https://www.mistartmap.info/) and were used to indicate when COVID-19 incidence exceeded 150 cases per million population per day. Dates when the “Stay Home, Stay Safe” order was implemented and when K-12 schools were closed were also obtained from the MI Safe Start Map.

Frequencies of survey responses were calculated and compared for each survey period. Serology data were combined into three serosurveillance periods: summer 2020, fall 2020-winter 2021, and spring 2021. Seroprevalence of antibodies against SARS-CoV-2 N protein was determined overall and by age group. Statistical analysis was performed using SAS (v9.4, SAS Institute Inc., Cary, NC) and R statistical software (v4.1.0; R core team, 2021).

## RESULTS

### Population, illness, and infection characteristics

A total of 1,606 individuals who were enrolled in the HIVE study contributed 23,502 person-months of follow-up during the study period. Participants included 740 adults and 866 children in 402 households. Most participants lived in households with 2 adults (n=1281, 79.8%) and 2 children (n=759, 47.3%) (Table 1). The number of adults in a household ranged from 1-6 and the number of children ranged from 0-7 (data not shown).

**Table 1.**
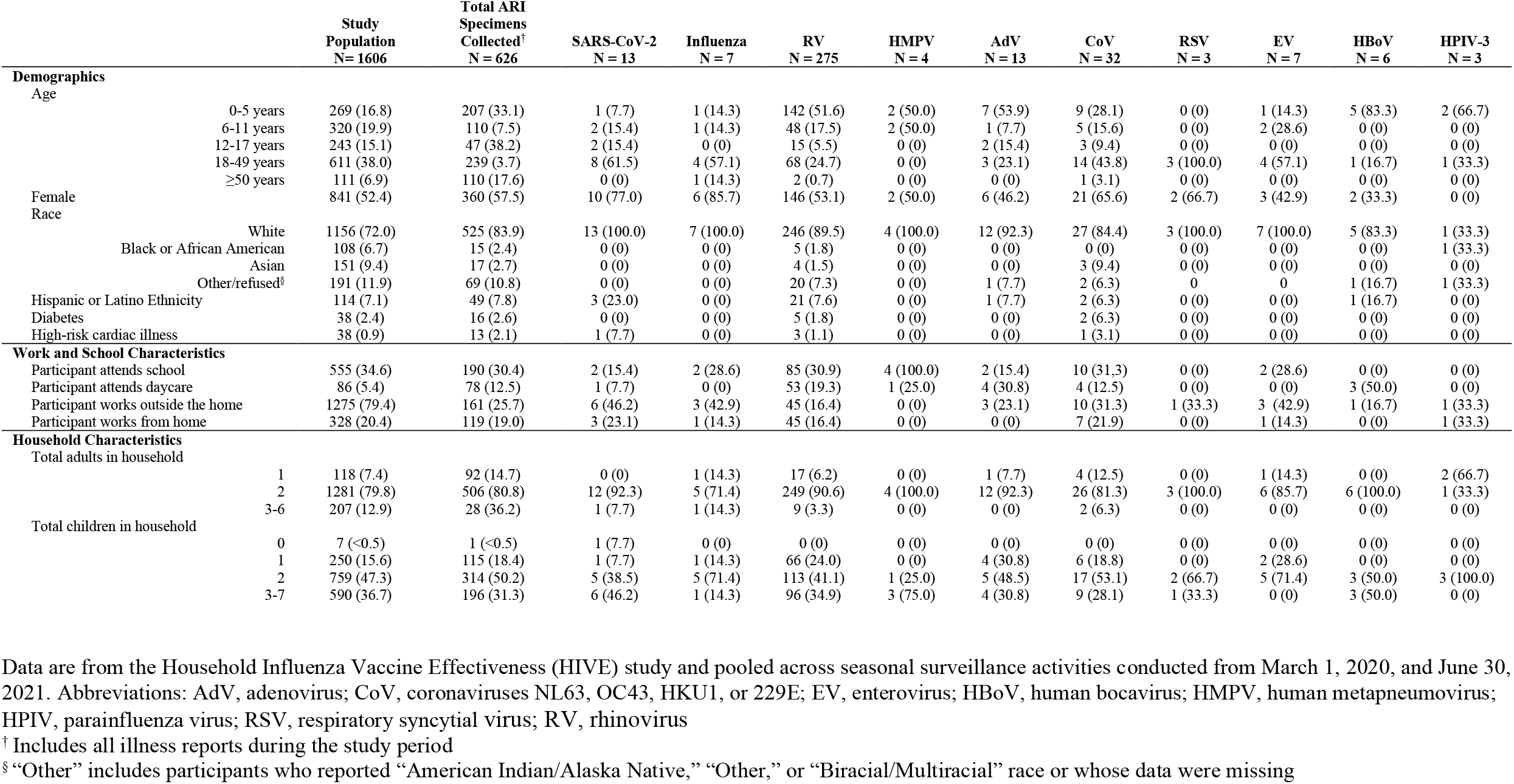
Demographic characteristics, illness reports, and infections among HIVE study participants illness — Michigan, March 2020–June 2021.

|  |  | Study<br>Population<br>N= 1606 | Total ARI<br>Specimens<br>Collected†<br>N = 626 | SARS-CoV-2<br>N = 13 | Influenza<br>N = 7 | RV<br>N = 275 | HMPV<br>N = 4 | AdV<br>N = 13 | CoV<br>N = 32 | RSV<br>N = 3 | EV<br>N = 7 | HBoV<br>N = 6 | HPIV-3<br>N = 3 |
| --- | --- | --- | --- | --- | --- | --- | --- | --- | --- | --- | --- | --- | --- |
| **Demographics** |  |  |  |  |  |  |  |  |  |  |  |  |  |
| Age |  |  |  |  |  |  |  |  |  |  |  |  |  |
|  | 0-5 years | 269 (16.8) | 207 (33.1) | 1 (7.7) | 1 (14.3) | 142 (51.6) | 2 (50.0) | 7 (53.9) | 9 (28.1) | 0 (0) | 1 (14.3) | 5 (83.3) | 2 (66.7) |
|  | 6-11 years | 320 (19.9) | 110 (7.5) | 2 (15.4) | 1 (14.3) | 48 (17.5) | 2 (50.0) | 1 (7.7) | 5 (15.6) | 0 (0) | 2 (28.6) | 0 (0) | 0 (0) |
|  | 12-17 years | 243 (15.1) | 47 (38.2) | 2 (15.4) | 0 (0) | 15 (5.5) | 0 (0) | 2 (15.4) | 3 (9.4) | 0 (0) | 0 (0) | 0 (0) | 0 (0) |
|  | 18-49 years | 611 (38.0) | 239 (3.7) | 8 (61.5) | 4 (57.1) | 68 (24.7) | 0 (0) | 3 (23.1) | 14 (43.8) | 3 (100.0) | 4 (57.1) | 1 (16.7) | 1 (33.3) |
|  | ≥50 years | 111 (6.9) | 110 (17.6) | 0 (0) | 1 (14.3) | 2 (0.7) | 0 (0) | 0 (0) | 1 (3.1) | 0 (0) | 0 (0) | 0 (0) | 0 (0) |
| Female |  | 841 (52.4) | 360 (57.5) | 10 (77.0) | 6 (85.7) | 146 (53.1) | 2 (50.0) | 6 (46.2) | 21 (65.6) | 2 (66.7) | 3 (42.9) | 2 (33.3) | 0 (0) |
| Race |  |  |  |  |  |  |  |  |  |  |  |  |  |
|  | White | 1156 (72.0) | 525 (83.9) | 13 (100.0) | 7 (100.0) | 246 (89.5) | 4 (100.0) | 12 (92.3) | 27 (84.4) | 3 (100.0) | 7 (100.0) | 5 (83.3) | 1 (33.3) |
|  | Black or African American | 108 (6.7) | 15 (2.4) | 0 (0) | 0 (0) | 5 (1.8) | 0 (0) | 0 (0) | 0 (0) | 0 (0) | 0 (0) | 0 (0) | 1 (33.3) |
|  | Asian | 151 (9.4) | 17 (2.7) | 0 (0) | 0 (0) | 4 (1.5) | 0 (0) | 0 (0) | 3 (9.4) | 0 (0) | 0 (0) | 0 (0) | 0 (0) |
|  | Other/refused§ | 191 (11.9) | 69 (10.8) | 0 (0) | 0 (0) | 20 (7.3) | 0 (0) | 1 (7.7) | 2 (6.3) | 0 | 0 | 1 (16.7) | 1 (33.3) |
| Hispanic or Latino Ethnicity |  | 114 (7.1) | 49 (7.8) | 3 (23.0) | 0 (0) | 21 (7.6) | 0 (0) | 1 (7.7) | 2 (6.3) | 0 (0) | 0 (0) | 1 (16.7) | 0 (0) |
| Diabetes |  | 38 (2.4) | 16 (2.6) | 0 (0) | 0 (0) | 5 (1.8) | 0 (0) | 0 (0) | 2 (6.3) | 0 (0) | 0 (0) | 0 (0) | 0 (0) |
| High-risk cardiac illness |  | 38 (0.9) | 13 (2.1) | 1 (7.7) | 0 (0) | 3 (1.1) | 0 (0) | 0 (0) | 1 (3.1) | 0 (0) | 0 (0) | 0 (0) | 0 (0) |
| **Work and School Characteristics** |  |  |  |  |  |  |  |  |  |  |  |  |  |
| Participant attends school |  | 555 (34.6) | 190 (30.4) | 2 (15.4) | 2 (28.6) | 85 (30.9) | 4 (100.0) | 2 (15.4) | 10 (31.3) | 0 (0) | 2 (28.6) | 0 (0) | 0 (0) |
| Participant attends daycare |  | 86 (5.4) | 78 (12.5) | 1 (7.7) | 0 (0) | 53 (19.3) | 1 (25.0) | 4 (30.8) | 4 (12.5) | 0 (0) | 0 (0) | 3 (50.0) | 0 (0) |
| Participant works outside the home |  | 1275 (79.4) | 161 (25.7) | 6 (46.2) | 3 (42.9) | 45 (16.4) | 0 (0) | 3 (23.1) | 10 (31.3) | 1 (33.3) | 3 (42.9) | 1 (16.7) | 1 (33.3) |
| Participant works from home |  | 328 (20.4) | 119 (19.0) | 3 (23.1) | 1 (14.3) | 45 (16.4) | 0 (0) | 0 (0) | 7 (21.9) | 0 (0) | 1 (14.3) | 0 (0) | 1 (33.3) |
| **Household Characteristics** |  |  |  |  |  |  |  |  |  |  |  |  |  |
| Total adults in household |  |  |  |  |  |  |  |  |  |  |  |  |  |
|  | 1 | 118 (7.4) | 92 (14.7) | 0 (0) | 1 (14.3) | 17 (6.2) | 0 (0) | 1 (7.7) | 4 (12.5) | 0 (0) | 1 (14.3) | 0 (0) | 2 (66.7) |
|  | 2 | 1281 (79.8) | 506 (80.8) | 12 (92.3) | 5 (71.4) | 249 (90.6) | 4 (100.0) | 12 (92.3) | 26 (81.3) | 3 (100.0) | 6 (85.7) | 6 (100.0) | 1 (33.3) |
|  | 3-6 | 207 (12.9) | 28 (36.2) | 1 (7.7) | 1 (14.3) | 9 (3.3) | 0 (0) | 0 (0) | 2 (6.3) | 0 (0) | 0 (0) | 0 (0) | 0 (0) |
| Total children in household |  |  |  |  |  |  |  |  |  |  |  |  |  |
|  | 0 | 7 (<0.5) | 1 (<0.5) | 1 (7.7) | 0 (0) | 0 (0) | 0 (0) | 0 (0) | 0 (0) | 0 (0) | 0 (0) | 0 (0) | 0 (0) |
|  | 1 | 250 (15.6) | 115 (18.4) | 1 (7.7) | 1 (14.3) | 66 (24.0) | 0 (0) | 4 (30.8) | 6 (18.8) | 0 (0) | 2 (28.6) | 0 (0) | 0 (0) |
|  | 2 | 759 (47.3) | 314 (50.2) | 5 (38.5) | 5 (71.4) | 113 (41.1) | 1 (25.0) | 5 (48.5) | 17 (53.1) | 2 (66.7) | 5 (71.4) | 3 (50.0) | 3 (100.0) |
|  | 3-7 | 590 (36.7) | 196 (31.3) | 6 (46.2) | 1 (14.3) | 96 (34.9) | 3 (75.0) | 4 (30.8) | 9 (28.1) | 1 (33.3) | 0 (0) | 3 (50.0) | 0 (0) |

Data are from the Household Influenza Vaccine Effectiveness (HIVE) study and pooled across seasonal surveillance activities conducted from March 1, 2020, and June 30, 2021. Abbreviations: AdV, adenovirus; CoV, coronaviruses NL63, OC43, HKU1, or 229E; EV, enterovirus; HBoV, human bocavirus; HMPV, human metapneumovirus; HPIV, parainfluenza virus; RSV, respiratory syncytial virus; RV, rhinovirus
† Includes all illness reports during the study period
§ “Other” includes participants who reported “American Indian/Alaska Native,” “Other,” or “Biracial/Multiracial” race or whose data were missing

A total of 772 ARI episodes from 437 individuals were reported between March 1, 2020, and June 30, 2021; 162 individuals reported ≥1 ARI episode (range: 1-11). The IR of ARI reports for the study period was 39.4 (95% CI: 36.7, 42.3) per 100 person-years. By comparison, the IR of ARI reports from a period of similar length before the pandemic (March 1, 2016, to June 30, 2017) was 76.9 (95% CI: 72.4, 81.6) per 100 person-years. Thus, the IRR of the incidence of ARI reports during the study period to the pre-pandemic period was 0.5 (95% CI: 0.5, 0.6, p<0.001).

Among 626 specimens collected at the time of ARI symptom onset, 343 tested positive for ≥1 respiratory virus. The IR of test-positive ARI specimens was 17.5 (95% CI: 15.7, 19.5) per 100 person-years. The IRR of the incidence of test-positive ARI specimens for the study period and the pre-pandemic comparison period was 0.3 (95% CI: 0.3, 0.4; p<0.001). ARI incidence declined immediately following statewide K-12 school closures in Michigan in mid-March 2020; incidence increased in September 2020 with the start of the 2020-2021 school year (Figure 1). Percent positivity for ≥1 respiratory virus from September 2020 through the end of the study was 60.7%.

**Figure 1.**
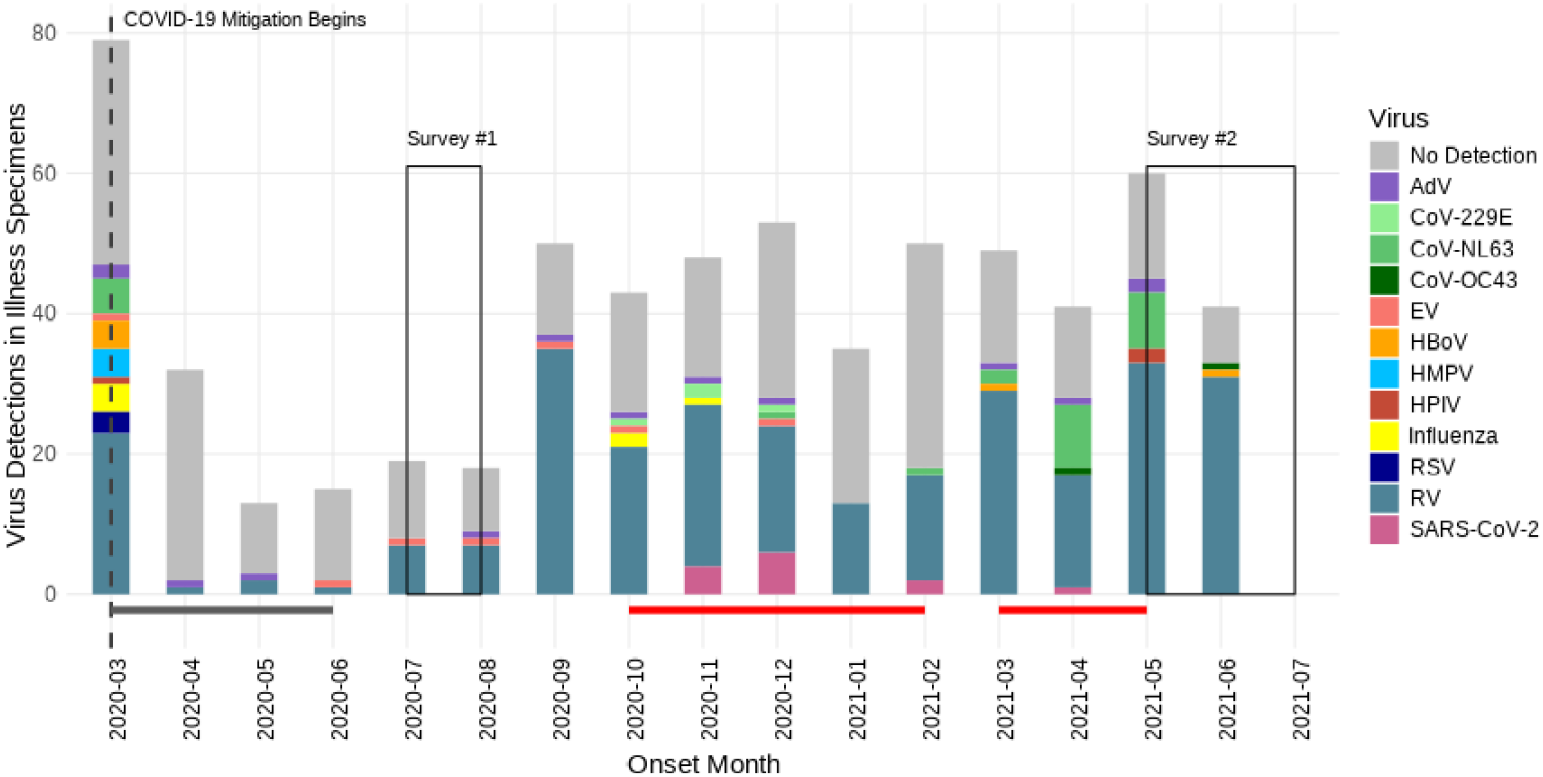
Epidemic curve of respiratory viruses from March 1, 2020, to June 30, 2021. Case counts are presented in a histogram with colors corresponding to specific agents. Illness reports are represented in the grey histogram behind case counts. Onset dates represent the date of illness onset according to MMWR week. The lines below the histogram represent the Stay Home Stay Safe period (dark grey line) and periods when COVID-19 cases were >150 million per person in Washtenaw County, MI (red line). *No Detection* indicates ARI reports for which a specimen was collected but all viral tests were negative. Abbreviations: AdV, adenovirus; CoV, coronavirus; EV, enterovirus; HBoV, human bocavirus; HMPV, human metapneumovirus; HPIV, parainfluenza virus; RSV, respiratory syncytial virus; RV, rhinovirus.

Rhinovirus, which was detected each month of the study period, was the most frequently detected virus (n=275, 80.2% of viruses detected), with 43.6% of all specimens testing positive for RV during the study period. The IR for ARI specimens that tested positive for RV was 14.0 (95% CI: 12.4, 15.8) per 100 person-years. The IRR of RV incidence during the study period and the pre-pandemic comparison period was 0.5 (95% CI: 0.5, 0.6; p<0.001).

Seasonal coronavirus NL63 followed RV in detection frequency (n=26, 7.6% of viruses detected), with 4.2% of all the specimens testing positive (Table 1). Influenza was detected in 7 ARI specimens (2.0% of positive tests), and SARS-CoV-2 was detected in 13 (3.8%). HPIV-3 was detected in 3 specimens (0.9% of positive tests). No participants reported ARI caused by coronavirus HKU1, but OC43, NL63, and 229E were identified between November 2020 and May 2021. RSV was detected only before the K-12 school closure. Of the positive specimens, 23 (6.7%) had co-detections of multiple pathogens. The most common coinfection during the study period was RV with AdV (n=6/23, 26.1%); there was one NL63 with SARS-CoV-2 coinfection observed (Table 1).

The age group which contributed the most ARI reports and accompanying specimens (n=239, 38.2%) was 18 to 49-year-olds (Table 1). Viral detection was more frequent in specimens from female participants (n=189, 55.1%) than male participants (n= 154, 44.9%). Positive specimens were most common among participants aged ≤5 years (n=161, 46.9%), followed by specimens among those 18 to 49 years of age (n=99, 28.9%).

### Mitigation Survey

A total of 326 adult participants from 211 households participated in the first survey (July-August 2020). A total of 89.8% (n=290) of respondents reported always wearing a face mask or cloth covering when in public; 69.6% (n=227) reported always avoiding gatherings of

≥10 people; and 82.7% (n=267) reported always covering coughs and sneezes (Table 2). Additionally, respondents reported greater handwashing frequency after being in public (n=277, 85.2%). These survey data were collected during the first summer of the COVID-19 pandemic when masking and social distancing requirements were strict and ARI reports were low (Figure 1).

**Table 2.**
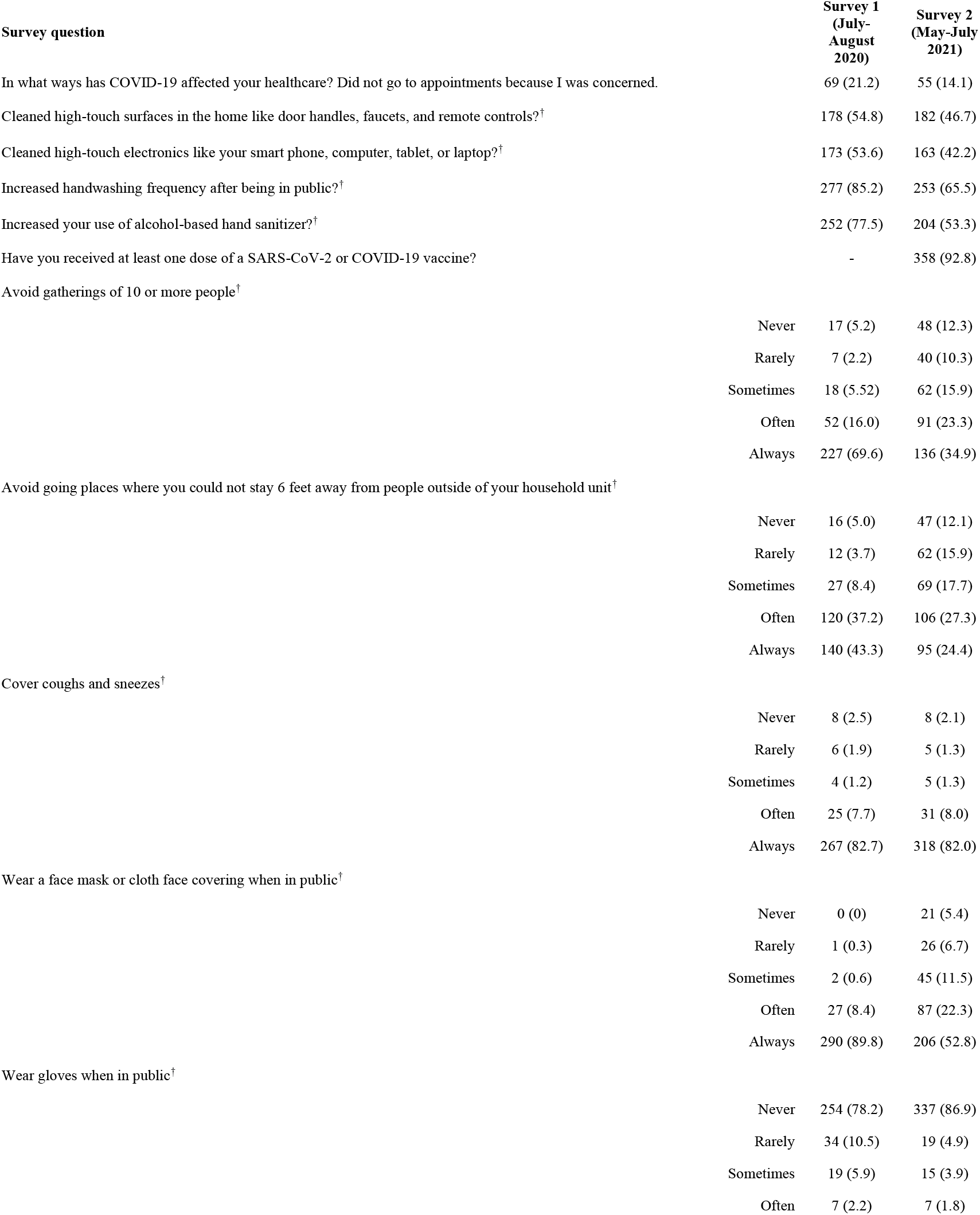
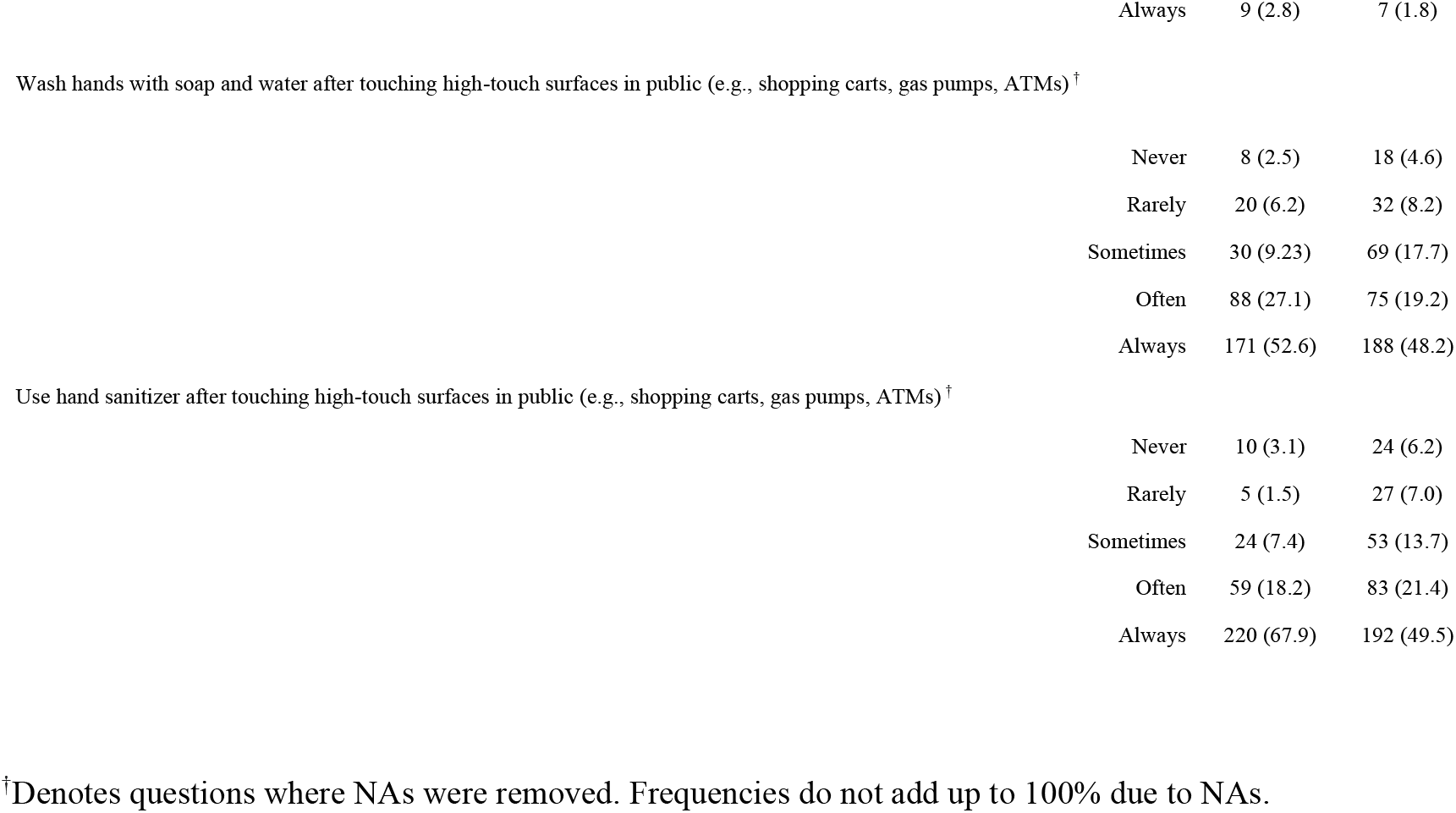
Mitigation survey results from HIVE participants.

A total of 390 participants from 262 households completed the second survey (May-July 2021)—including 234 people from 165 households who participated in both surveys. Approximately 40% fewer participants (n= 206, 52.8%) in the second survey reported always wearing a face mask or cloth covering when in public than relative to the first survey (Table 2). The percentage of participants who reported always avoiding gatherings of ≥10 people dropped from 69.6% in the first to 34.9% (n=136) in the second survey, but many participants continued to report always washing their hands with soap and water after touching high-touch surfaces in public (n=188, 48.2%) and always covering coughs and sneezes (n=318, 82.0%). It is important to consider that, by the time of survey administration, 92.8% (n=358) of the second survey’s participants had received ≥1 dose of a COVID-19 vaccine and mitigation measures had been relaxed. We noted an increase in ARI reports and infections—corresponding to the timing of the second survey—when diminished adherence to mitigation practices was reported (Figure 1).

### SARS-CoV-2 Serology

Blood specimens were collected from 409 subjects from March 1, 2020, through April 30, 2021, in three serosurveillance periods (7/17/2020-8/31/2020 [n=265], 9/1/2020-2/3/2021 [n=269], and 3/9/2021-4/30/2021 [n=62]). Of the specimens, 98 were from children ≤12 years of age (who were ineligible for vaccination during the study period). Overall, 29 (7.1%) specimens tested seropositive for antibodies against the SARS-CoV-2 N protein, indicating prior SARS-CoV-2 infection. There were 14 (5.3% of 265) collected during summer 2020; 8 (3.0% of 269) during fall-winter 2020; and 7 (11.3% of 62) in spring 2021. Only 2 seropositive specimens (7% of 29) were from individuals less than 12 years old. Among 29 individuals who tested seropositive, 2 tested N antibody-seronegative during a subsequent interval (106 and 284 days after the initial seropositive specimen, respectively). Of the 29 seropositive individuals, 13 reported a total of 26 ARI episodes during the March 1, 2020, to June 30, 2021, period; 3 (11.5%) illness specimens were positive for SARS-CoV-2 by RT-PCR. The remaining individuals with detected antibodies did not have a SARS-CoV-2 infection identified by study surveillance.

## DISCUSSION

The presented study, which began before the COVID-19 pandemic, offered an opportunity to observe the occurrence and patterns of circulating respiratory viruses during the first year of the pandemic. Rhinovirus infections were detected throughout the study period. Seasonal coronaviruses were detected in the winter months; only sporadic influenza, RSV, and SARS-CoV-2 detections were made during this time in the study area. Surveillance reports of ARI and detected viral infections within study participants increased after the statewide “Stay Home, Stay Safe” mandate ended. In contrast, ARI incidence in the cohort was higher between fall 2020 and winter 2021 (i.e., when respondents reported less adherence to mitigation practices relative to summer of 2020). Seroprevalence of SARS-CoV-2 antibodies was low throughout the study period.

Many of the non-pharmaceutical interventions deployed worldwide at the beginning of the COVID-19 pandemic were originally developed as a strategy for early reduction of peak transmission levels during an influenza pandemic, so the reduction in the circulation of many non-SARS-CoV-2 respiratory viruses was concurrent to the adoption of mitigation and continued high levels of influenza vaccination in the study participants was not surprising.^15–18^ However, as influenza pandemics pass through communities in a matter of months, the maintenance of these measures over longer periods during multiple waves of COVID-19 may have disrupted the seasonality of other respiratory viruses to a degree never before seen.^19–22^

The mechanism for the disruption of seasonal respiratory viruses by SARS-CoV-2 circulation—whether via non-pharmaceutical mitigation or an interference effect—remains unclear. One hypothetical mechanism is a biological interference between SARS-CoV-2 and other respiratory viruses, which has been observed in animal models and is presumed to occur on the population scale.^23–27^ Crucially, it is likely that social and behavioral factors have played a significant role in these patterns, particularly the coinciding occurrence of such stark mitigation measures during this time (e.g., school and business closures, and regional travel restrictions). These major alterations in contact patterns must be considered in conjunction with any biological hypotheses.

Rhinovirus was the most common respiratory virus detected in participant specimens and was detected throughout the study. Rhinovirus detection decreased noticeably when K-12 schools closed in March 2020 and increased in May 2021 when public health mitigation measures were relaxed in Michigan. Kitanovski and colleagues^28^ reported that the prevalence of RV in a cohort of ambulatory and hospitalized patients in Germany was negatively correlated with the stringency of COVID-19 precautions. Our findings reflect this trend and suggest that, though RV was detected throughout the study period, its detection was impacted by changes in mitigation measure prescription and adherence. Specifically, detection decreased when mitigation measures were stringent and increased when mitigation measures were relaxed by way of statewide policies and as supported by surveys of individuals’ behavior.

During pandemic virus circulation, in addition to decreased infection rates due to public safety precautions, ARI transmission patterns may differ from normal seasonal patterns. Haddadin and colleagues^29^ recently described a multicenter study on ARIs in children during the COVID-19 pandemic and found a lower rate of ARI cases per week in most areas compared with prior years. However, temporal trends were roughly equivalent from 2016 to 2020. Notably, our prospective cohort study of mild-to-moderate illness found that the most highly seasonal viruses, influenza and RSV, were scarce, suggesting substantial changes in the transmission of these viruses. The incidence of ARI reports decreased by 50% during the study period compared to a pre-pandemic comparison period. Similarly, compared to the pre-pandemic comparison period, the incidence rate of test-positive ARI specimens decreased by 70% during the study period. Lastly, the incidence of RV during the study period was approximately 50% lower relative to the comparison period.

Supplemental serosurveillance identified SARS-CoV-2 infections that were not identified by surveillance using our study case definition of ARI. Ten of the 29 individuals with detected antibodies did not have SARS-CoV-2 detected during HIVE ARI surveillance, suggesting that only one-third of infections were detected even with active surveillance and swabbing. Asymptomatic infections, syndromic presentations that met criteria for swab collection, and/or underreporting by participants likely contributed. These results highlight the use of serology in addition to RT-PCR to capture additional infections in a cohort.^30,31^ Still, the relatively low frequency of incident SARS-CoV-2 infections suggests the effectiveness of non-pharmaceutical interventions in these households despite high COVID-19 incidence in the community during the study period. Furthermore, it is important to note that vaccine coverage in the HIVE population has historically been extremely high. This may also help to explain the adherence to and efficacy of non-pharmaceutical interventions in preventing SARS-CoV-2 infections.

This study has several limitations. Respiratory virus infections may have gone undetected by study assays if participants had viral loads below detectable limits at the time of collection. HIVE participants were asked to collect specimens as soon as possible after a reported illness to improve virus detection. ARI episodes in this cohort were mild and may not be representative of more severe illness; other studies of ARIs during the pandemic have measured disease severity among children seeking medical attention at the hospital, among whom respiratory virus positivity may differ.^32,33^ Generalizability to other populations is limited as participants in the HIVE study may not be representative of the general population.

The incidence of ARI in the HIVE cohort declined during the pandemic concurrent with the widespread use of public health measures designed to reduce transmission of SARS-CoV-2; however, RV and seasonal coronaviruses continued to circulate. Prospective household studies such as HIVE that incorporate systematic, active case-finding and multi-pathogen surveillance are important contributors to our understanding of respiratory virus transmission in a community. Longitudinal cohorts capture illnesses that do not necessarily come to the attention of medical systems, and the milder illness events captured therein are critical to fully describe the variety of circulating respiratory viruses. These surveillance systems will continue to be a resource for the evaluation of mitigation strategies both during seasonal epidemics and in future pandemics.

## Data Availability

All data produced in the present study are available upon reasonable request to the authors and accompanying approval by the University of Michigan Institutional Review Board.

## Acknowledgments

We are grateful for the participants who contributed to the HIVE study.

---

**Table S1.** Coinfections detected among HIVE study participants.

| **Coinfection** | **Count (%)**<br>**n=23** |
| --- | --- |
| RV, ADV | 6 (26.1) |
| RV, HMPV | 2 (8.7) |
| RV, HBoV | 2 (8.7) |
| RV, SARS-CoV-2 | 3 (13.0) |
| RV, CoV 229E | 1 (4.3) |
| RV, CoV OC43 | 1 (4.3) |
| CoV NL63, EV | 1 (4.3) |
| CoV NL63, SARS-CoV-2 | 1 (4.3) |
| ADV, HBoV | 1 (4.3) |
| Influenza A, Influenza B | 1 (4.3) |
| Influenza A, RSV | 1 (4.3) |
| Influenza A, CoV 229E | 1 (4.3) |
| HPIV-3, HBoV | 1 (4.3) |
| RV, ADV, HBoV | 1 (4.3) |

Abbreviations: AdV, adenovirus; CoV, coronavirus; EV, enterovirus; HBoV, human bocavirus; HMPV, human metapneumovirus; HPIV, parainfluenza virus; RSV, respiratory syncytial virus.

